# Evaluation of height as a disease risk factor through a phenome-wide association study of genetically-predicted height

**DOI:** 10.1101/2021.08.29.21262793

**Authors:** Sridharan Raghavan, Jie Huang, Catherine Tcheandjieu, Jennifer E. Huffman, Elizabeth Litkowski, Yuk-Lam A. Ho, Haley Hunter-Zinck, Hongyu Zhao, Eirini Marouli, Kari E. North, the VA Million Veteran Program, Ethan Lange, Leslie A. Lange, Benjamin F. Voight, J. Michael Gaziano, Saiju Pyarajan, Elizabeth R. Hauser, Philip S. Tsao, Peter W. F. Wilson, Kyong-Mi Chang, Kelly Cho, Christopher J. O’Donnell, Yan V. Sun, Themistocles L. Assimes

**Affiliations:** Medicine Service, Veterans Affairs Eastern Colorado Health Care System, Aurora, CO, USA; Department of Medicine, University of Colorado Anschutz Medical Campus, Aurora, CO, USA; Department of Global Health, Peking University School of Public Health, Beijing, China; Veterans Affairs Palo Alto Health Care System, Palo Alto, CA, USA; Division of Cardiovascular Medicine, Department of Medicine, Stanford University School of Medicine, Stanford, CA, USA; Veterans Affairs Boston Healthcare System, Boston, MA, USA; Veterans Affairs Connecticut Healthcare System, West Haven, CT, USA; Department of Biostatistics, Yale University School of Public Health, New Haven, CT, USA; William Harvey Research Institute, Barts and The London School of Medicine and Dentistry, Queen Mary University of London, London, UK; Department of Epidemiology, Gillings School of Public Health, University of North Carolina, Chapel Hill, NC, USA; Corporal Michael J. Crescenz Veterans Affairs Medical Center, Philadelphia, PA, USA; Department of Genetics, University of Pennsylvania Perelman School of Medicine, Philadelphia, PA, USA; Department of Systems Pharmacology and Translational Therapeutics, University of Pennsylvania Perelman School of Medicine, Philadelphia, PA, USA; Institute of Translational Medicine, University of Pennsylvania Perelman School of Medicine, Philadelphia, PA, USA; Department of Medicine, Brigham and Women’s Hospital, Harvard Medical School, Boston, MA, USA; Cooperative Studies Program Epidemiology Center- Durham, Durham Veterans Affairs Health Care System, Durham, NC, USA; Department of Biostatistics and Bioinformatics, Duke University Medical Center, Durham, NC, USA; Atlanta Veterans Affairs Medical Center, Atlanta, GA, USA; Division of Cardiology, Emory University School of Medicine, Atlanta, GA, USA; Department of Medicine, University of Pennsylvania Perelman School of Medicine, Philadelphia, PA, USA; Department of Epidemiology, Emory University Rollins School of Public Health, Atlanta, GA, USA

**Author notes:** Corresponding author: Sridharan Raghavan, MD, PhD, Rocky Mountain Regional VA Medical Center, Medicine Service (111), 1700 North Wheeling Street, Aurora, CO 80045. Equally contributing authors (Sridharan Raghavan, Jie Huang).

## Abstract

**Background:** Height has been associated with many clinical traits but whether such associations are causal versus secondary to confounding remains unclear in many cases. To systematically examine this question, we performed a Mendelian Randomization-Phenome-wide association study (MR-PheWAS) using clinical and genetic data from a national healthcare system biobank.

**Methods and Findings:** Analyses were performed using data from the US Veterans Affairs (VA) Million Veteran Program in non-Hispanic White (EA, n=222,300) and non-Hispanic Black (AA, n=58,151) adults in the US. We estimated height genetic risk based on 3290 height-associated variants from a recent European-ancestry genome-wide meta-analysis. We compared associations of measured and genetically-predicted height with phenome-wide traits derived from the VA electronic health record, adjusting for age, sex, and genetic principal components. We found 345 clinical traits associated with measured height in EA and an additional 17 in AA. Of these, 127 were associated with genetically-predicted height at phenome-wide significance in EA and 2 in AA. These associations were largely independent from body mass index. We confirmed several previously described MR associations between height and cardiovascular disease traits such as hypertension, hyperlipidemia, coronary heart disease (CHD), and atrial fibrillation, and further uncovered MR associations with venous circulatory disorders and peripheral neuropathy. As a number of traits associated with genetically-predicted height frequently co-occur with diabetes mellitus and/or CHD, we evaluated effect modification by diabetes and CHD status of genetically-predicted height associations with risk factors for and complications of diabetes and CHD. We found modification of effects of MR associations by diabetes for skin and bone infections and by CHD status for atrial fibrillation/flutter.

**Conclusions:** We conclude that height may be an unrecognized but biologically plausible risk factor for several common conditions in adults. However, more studies are needed to reliably exclude horizontal pleiotropy as a driving force behind at least some of the MR associations observed in this study.

## Introduction

Height is not typically considered a disease risk factor but has nevertheless been associated with numerous diseases^1-5^. Such epidemiologic associations of height with disease endpoints are susceptible to confounding as adult height is also influenced by environmental factors, including nutrition, socioeconomic status, and demographic factors^1,6-9^. The high heritability of height coupled with recent advances in understanding its genetic basis^10-12^ now make it possible to use genetic tools to elucidate pathophysiologic relationships between height and clinical traits.

Mendelian randomization (MR) is an instrumental variable approach that utilizes genetic instruments for exposures of interest under the assumption that genotype is less susceptible to confounding than measured exposures^13^. Indeed, MR has recently been used to address unmeasured confounding and to estimate causal effects of height on several candidate traits of interest, including coronary heart disease (CHD), lipid levels, atrial fibrillation, and certain cancers^6,14-20^. The largest of the MR studies examined height associations with 50 traits using data from the UK Biobank and 691 height-associated genetic variants from a European-ancestry genome-wide association study (GWAS) meta-analysis^11,15^. Twelve of the 50 traits studied had genetic evidence supporting an association with height: risk-lowering associations of taller stature with heart disease, hypertension, diaphragmatic hernia, and gastroesophageal reflux disease, and risk-increasing associations of height with atrial fibrillation, venous thromboembolic events, hip fracture, intervertebral disc disease, vasculitis, and cancer (all-cause as well as breast and colorectal cancers specifically)^15^. Thus, the prior work expands the genetic evidence supporting height associations beyond cardiovascular disease and cancer to gastrointestinal, musculoskeletal, and rheumatologic diseases.

While the prior MR studies of height have tested hypotheses based on previously described epidemiologic associations, MR methods have also been combined with phenome-wide association studies (MR-PheWAS) to identify novel or hypothesis-generating associations^21^. For example, using a genetic instrument for body mass index (BMI) and phenotype data from the UK Biobank, a recent study evaluated genetic evidence for associations of BMI with nearly 20,000 independent traits and identified novel associations with psychiatric traits related to nervousness^22^. A similar comprehensive or phenome-wide evaluation of clinical traits associated with measured and genetically-predicted height could elucidate the full scope of diseases associated with height as a risk or protective factor. To that end, we performed an MR-PheWAS of height in the multiethnic US Department of Veterans Affairs (VA) Million Veteran Program (MVP).

## Methods

### Study Participants

The design of the MVP has been previously described^23,24^. Briefly, participants were recruited from over 60 VA medical centers nationwide starting in 2011. Enrolled Veterans provided a blood sample for banking from which DNA was extracted for genotyping. Phenotype data for the MVP is derived from the VA electronic health record (EHR), integrating inpatient and outpatient International Classification of Diseases (ICD 9/10) diagnosis and procedure codes, Current Procedural Terminology (CPT) procedure codes, clinical laboratory measurements, medications, and reports of diagnostic imaging modalities into a clinical research database. Ethical oversight and human subjects research protocol approval for the MVP were provided by the VA Central Institutional Review Board in accordance with the principles outlined in the Declaration of Helsinki.

### Anthropometric traits

Height and body mass index (BMI) were based on EHR data from clinical examinations available for participants from 2003 through 2018. For height, we used the average of all measurements available for a participant, excluding height measurements that were >3 inches above or below the average for each participant. Individuals with extreme average heights (≤50 inches or ≥100 inches) were also excluded. Average height was then converted to centimeters for all subsequent analyses. BMI was calculated as the weight (in kilograms) divided by the height (in meters) squared. We calculated the average BMI using all measurements occurring within 1.5 years before to 1.5 years after the date of MVP enrollment excluding weight measurements >60 pounds from the average of each participant.

### Genetic Data

Genotyping, quality control procedures, and imputation in the MVP has been described previously^23^. Briefly, DNA extracted from blood was genotyped using a customized Affymetrix Axiom® biobank array (MVP 1.0 Genotyping Array) with content enriched for common and rare genetic variants of clinical significance. Duplicate samples and samples with excess heterozygosity, with >2.5% of missing genotype calls, or with sex-gender discordance were excluded. In cases of pairs of related individuals, one individual from each pair was removed. Variants with poor calling or with allele frequencies discrepant from 1000 Genomes Project^25^ reference data were excluded. Genotypes from the 1000 Genomes Project phase 3 reference panel were imputed into MVP as described previously^26^. After imputation, variant level quality control was performed with the following exclusion thresholds: ancestry-specific Hardy-Weinberg equilibrium^27^ *p-*value <1×10^−20^, posterior call probability < 0.9, imputation quality <0.3, minor allele frequency (MAF) < 0.0003, call rate < 97.5% for common variants (MAF > 1%), and call rate < 99% for rare variants (MAF < 1%). Finally, we performed global and ancestry-specific principal component analysis (PCA) using the flashPCA software.

### Genetic risk score for height

We used 3,290 independent, genome-wide significant variants and their beta coefficients from a previously published GWAS in individuals of European ancestry^10^ with no overlap with MVP to build a genetic risk score (GRS) for height. The weighted genetic risk score for height was the sum of the number of height-raising alleles in each individual, weighting each variant by the beta estimate for its effect on height form the published source GWAS^10^.

### PheWAS phenotypes

Case/control status of MVP participants for 1,813 phecodes^28,29^ was assigned using EHR data, based on data available at the time of enrollment into the MVP. Analyses were limited to phecodes with at least 200 case and 200 non-case participants.

### PheWAS and MR-PheWAS of height

Race/ethnicity group was assigned using the Harmonized Ancestry and Race/Ethnicity (HARE) algorithm, which integrates self-identified race/ethnicity with genetically inferred ancestry^30^. Using HARE, we classified individuals as non-Hispanic White (EA), non-Hispanic Black (AA), and Hispanic-American (HA). PheWAS analyses were performed in a total of 222,300 EA and 58,151 AA participants with non-missing measured height, height GRS, and EHR phenotype data. To estimate associations of height with EHR traits, we performed race/ethnicity-stratified logistic regression of phecodes as the dependent variable and height Z-scores as the independent variable, adjusting for age at MVP enrollment, sex, and 10 race/ethnicity-specific PCs. In the MR-PheWAS, associations of EHR traits with genetically-estimated height were performed using a two-stage least squares regression^13^. First, we performed race/ethnicity-stratified linear regression of measured height (dependent variable) on the height GRS, adjusting for 10 race/ethnicity-specific PCs. Next, we estimated genetically-predicted height for each participant using the first-step regression equation, and subsequently standardized genetically-predicted height to the standard deviation of measured height. Thus, a one unit change in the standardized genetically-predicted height corresponds to a one unit change in the height Z-score. We then used the R *PheWAS* package^31^ to perform a race/ethnicity-stratified MR-PheWAS which used logistic regression to estimate associations of phecodes as the dependent variables and standardized genetically-predicted height as the independent variable, adjusting for age at enrollment in MVP, sex, and 10 race/ethnicity-specific PCs. Results of the MR-PheWAS, therefore, indicate the odds ratio (OR) for a phecode per unit change in genetically-predicted height, scaled to the standard deviation of measured height in our sample to allow direct comparability between the PheWAS and MR-PheWAS results. The number of phecodes included in the analysis of EA individuals was 1,379 and of AA individuals was 1,026, resulting in corrected thresholds for significance of *p*<3.6×10^−5^ (0.05 / 1,379) for EA and *p*<4.8×10^−5^ (0.05 / 1,026) for AA analyses. We reported nominally-significant association (*p*<0.05) results of the PheWAS in AA given the much smaller sample size and the fact that the GRS weights were derived from a European-ancestry GWAS.

We tested concordance in direction of effect of phenome-wide significant traits for height-trait and genetically-predicted height-trait associations between EA and AA analyses using chi-square tests. For each of these pairwise comparisons, we also compared the magnitudes of beta coefficients for phenome-wide significant traits using the slope of the linear regression of beta coefficients arising from each pair of analyses being compared (e.g., height-trait associations between EA and AA).

### Secondary analyses

We performed several secondary analyses motivated by the results of the PheWAS and previously known relationships between height and clinical characteristics. As height and body mass index (BMI; calculated as weight in kilograms divided by height in meters squared) are correlated, we ran an additional model with BMI as an added covariate to determine the extent to which genetically-predicted height associations with phecodes could be confounded by BMI. In addition, we repeated the PheWAS stratified by diabetes and CHD status to address two potential issues. First, differential recognition and documentation in the EHR of conditions in individuals with diabetes or CHD due to more frequent or intense clinical care could induce false positive associations in the MR-PheWAS. This is particularly of concern for CHD-associated conditions given the previously described associations of height with CHD. Second, the stratified analyses might reveal differential associations of genetically-predicted height with conditions that could suggest synergistic effects of diabetes or CHD with height on those conditions. As some of the traits most strongly associated with genetically-predicted height are potentially complications of or correlated with diabetes or CHD, we selected these common conditions that are more highly prevalent among Veterans than in the general population^32-34^ for the stratified analyses. For diabetes, stratification was based on the presence/absence of phecodes related to diabetes and diabetes-related conditions: 250, 250.1, 250.11, 250.12, 250.13, 250.14, 250.15, 250.2, 250.21, 250.22, 250.23, 250.24, 250.25, 250.6, and 249. For CHD, stratification was based on the presence/absence of phecodes related to coronary atherosclerosis: 411, 411.1, 411.2, 411.3, 411.4, 411.41, 411.8, and 411.9.

### GWAS

Lastly, we performed a GWAS of height in the MVP sample for two reasons. First, we wanted to compare the genetic associations with height in MVP of the variants used to generate the height GRS across non-European ancestries. Second, we wanted to leverage the multi-ethnic MVP sample containing a substantial sample size of non-EA participants to perform a trans-ethnic meta-analysis of height to determine if inclusion of diverse ancestries yielded novel genetic associations with height. GWAS of height in the MVP cohort was examined separately among EA (N=235,398), AA (N=63,898), and HA (N=24,497) participants based on the HARE algorithm for classifying race/ethnicity^30^. For each race/ethnicity group, height was stratified by sex and adjusted for age, age^2^, and the top ten genotype-derived PCs in a linear regression model. The resulting residuals were transformed to approximate normality using inverse normal scores.

Imputed and directly measured genetic variants were tested for association with the inverse normal transformed residuals of height through linear regression assuming an additive genetic model. We performed inverse-variance weighted fixed-effects meta-analysis using METAL^35^. For the minority ancestries in our sample, we meta-analyzed the GWAS results from MVP AA and HA participants, and we performed a trans-ethnic meta-analysis within MVP of EA, AA, and HA results. GWAS results were summarized using FUMA (http://fuma.ctglab.nl/), a platform that annotates, prioritizes, visualizes and interprets GWAS results^36^. Independent, genome-wide significant SNPs were defined as those with p < 5×10^−8^ and with LD r^2^ < 0.6 with each other. SNPs with p < 0.05 were grouped into a genomic locus if they were linked at r^2^ ≥ 0.6 or were physically close (distance < 500kb). Lead SNPs were defined within each locus if they were independent (r^2^ < 0.1) and genome-wide significant (p < 5×10^−8^). Novel loci were defined as those with genome-wide significance (p < 5×10^−8^) and a distance > 500kb from previously published variants in European and African ancestries^10,37^. All GWAS summary statistics will be deposited in the database of genotypes and phenotypes (dbGaP) at the time of publication.

To quantify the transferability of the beta coefficients from the European-ancestry GWAS used to build the height GRS to the MVP sample, we tested directional concordance of the association with height of the 3,290 variants included in the GRS using a chi-square test. Additionally, we calculated the standardized Z’^38,39^ for the effect size of variant associations with height in MVP and in the published GWAS and estimated the slope of the regression line between effect sizes in the two samples with a slope of 1 in the regression indicating equivalent effect sizes in both samples.

## Results

### Study participant characteristics

Among 323,793 MVP participants with both genetic data and height measurements, there were 73% of EA (n=235,398), 20% of AA (n=63,898), and 7.6% of HA (n=24,497) race/ethnicity. The MVP participants were predominantly men (91.6%) with a mean height of 176 cm and mean BMI of 30.1 kg/m^2^ (**Table 1**). AA and EA participants were similar with regard to sex, BMI, and height, but AA participants were younger on average (57.7 years versus 64.2 years). HA participants were younger (mean age 55.7 years) and shorter (172 cm) than AA and EA counterparts (**Table 1**). Of these, 280,451 individuals (222,300 EA and 58,151 AA) were included in the PheWAS analysis.

**Table 1.**
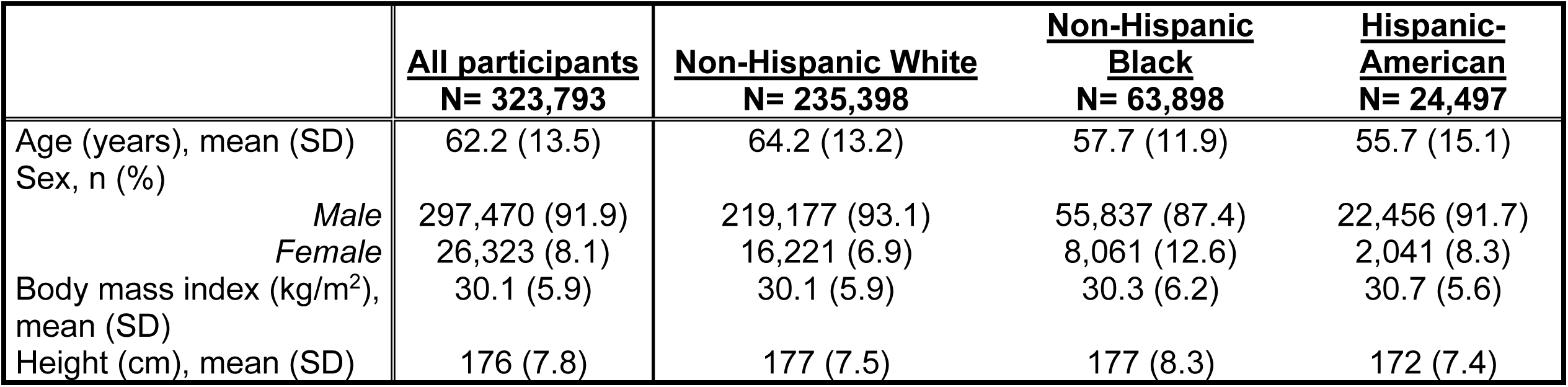
Study participant characteristics.

|  | **<u>All participants</u>**<br>**N= 323,793** | **<u>Non-Hispanic White</u>**<br>**N= 235,398** | **<u>Non-Hispanic Black</u>**<br>**N= 63,898** | **<u>Hispanic-American</u>**<br>**N= 24,497** |
| --- | --- | --- | --- | --- |
| Age (years), mean (SD) | 62.2 (13.5) | 64.2 (13.2) | 57.7 (11.9) | 55.7 (15.1) |
| Sex, n (%) |  |  |  |  |
| *Male* | 297,470 (91.9) | 219,177 (93.1) | 55,837 (87.4) | 22,456 (91.7) |
| *Female* | 26,323 (8.1) | 16,221 (6.9) | 8,061 (12.6) | 2,041 (8.3) |
| Body mass index (kg/m<sup>2</sup>), mean (SD) | 30.1 (5.9) | 30.1 (5.9) | 30.3 (6.2) | 30.7 (5.6) |
| Height (cm), mean (SD) | 176 (7.8) | 177 (7.5) | 177 (8.3) | 172 (7.4) |

### Transferability of genetic associations with height to the MVP population

The GRS based on 3,290 SNPs independently associated with height in a prior European-ancestry meta-analysis that did not include MVP explained 18% of height variation in EA and 4.8% of height variation in AA in MVP. Of the 3,290 variants examined, 89% had concordant direction of effect between the discovery study and MVP EA (*p* < 2×10^−16^) and 72% had concordant direction of effect between the discovery study and MVP AA (*p* < 2×10^−16^). The effect sizes for associations between the 3,290 SNPs and height were well-correlated between the discovery study and MVP EA and AA with a best-fit line slope of 0.87 [95% CI 0.84, 0.90] in EA and of 0.78 [0.73, 0.82] in AA (**Supplemental Figure 1**).

### Association of height and genetically-predicted height with clinical traits

A total of 345 traits were associated with measured height at phenome-wide significance (*p* < 3.6×10^−5^) among EA individuals, 132 of which were phenome-wide significant (*p* < 4.8×10^−5^) in AA individuals as well (**Figure 1A**). An additional 17 traits were phenome-wide significant in AA individuals but not in EA individuals (**Supplemental Tables 1 and 2**). The effect estimates for height-trait associations that reached phenome-wide significance in at least one race/ethnicity group were well correlated (r = 0.87, *p* = 1.1×10^−114^) between EA and AA individuals with the regression line of best fit between the two having a slope of 0.93 [95% CI 0.89, 0.98] (**Figure 1A**). A total of 335 out of 362 (213 in EA only, 132 in both AA and EA, and 17 in AA alone) phenome-wide significant height-trait associations were directionally concordant across the two ancestries (*p* < 2×10^−16^). A total of 142 traits were associated with genetically-predicted height at phenome-wide significance among EA individuals, 2 of which were phenome-wide significant in AA individuals as well, and another 46 of which were nominally significant (*p* < 0.05) in AA individuals (**Figure 1B, Supplemental Tables 3 and 4**). The effect estimates for phenome-wide significant associations of clinical traits with genetically-predicted height were reasonably correlated between EA and AA individuals (r=0.62, *p* = 1.5×10^−16^; best fit line slope 0.78 [95% CI 0.66, 0.90]) and improved when limited to traits that were at least nominally significant in AA individuals (r = 0.90, *p* = 4.2×10^−18^; best fit line slope 0.88 [95% CI 0.81, 0.96]) (**Figure 1B**). We observed strong directional concordance of phenome-wide significant trait associations with genetically-predicted height across the two ancestries (124 out of 142 traits; *p* = 4×10^−16^).

**Figure 1.**
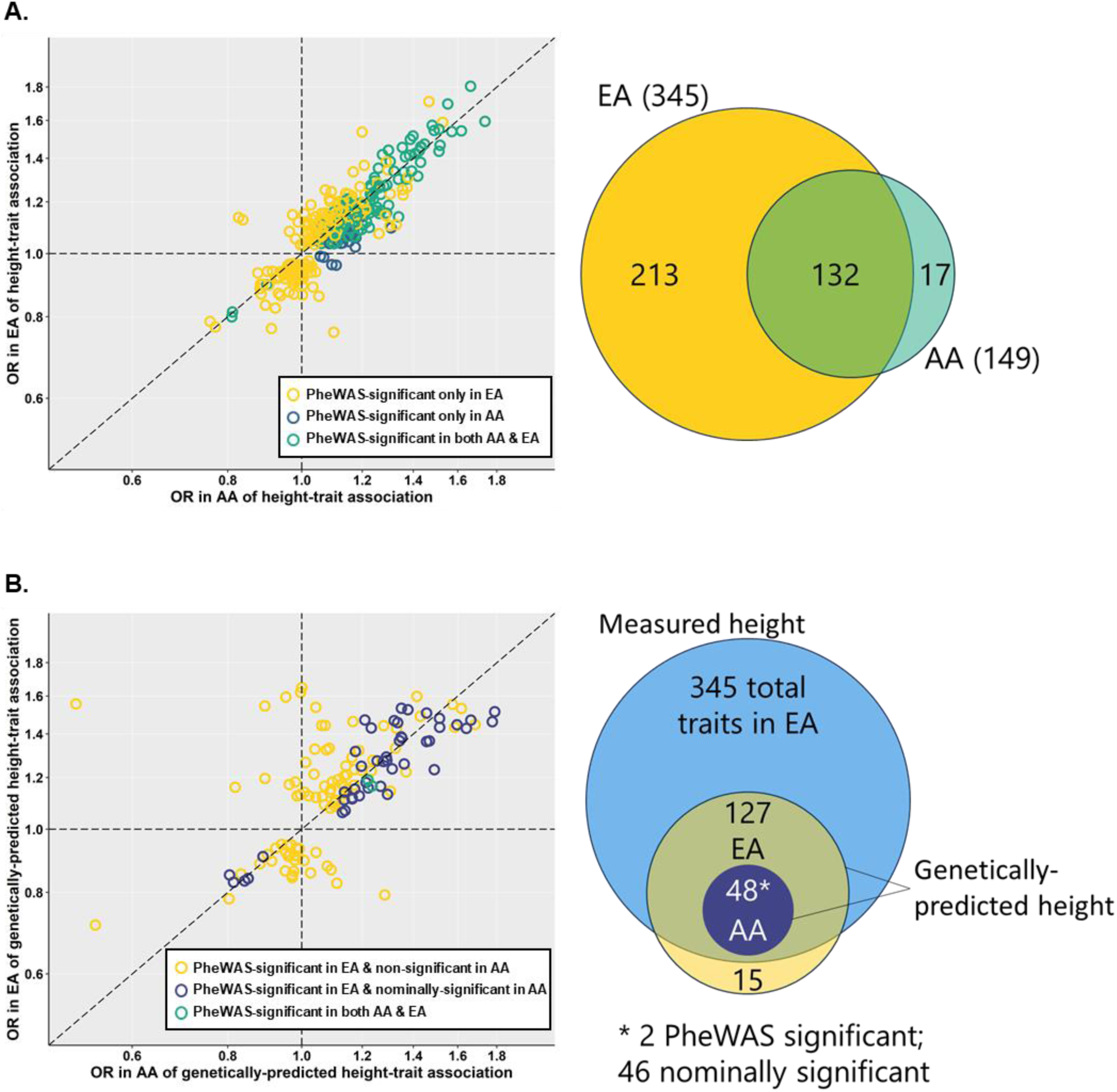
Comparison of number of associations and effect sizes of measured height (A) and genetically-predicted height (B) between non-Hispanic White and non-Hispanic Black individuals. Associations of measured and genetically-predicted height with phecodes represented as odds ratios (OR) in non-Hispanic Black (AA) and non-Hispanic White (EA) MVP participants. Whether associations exceeded phenome-wide significance threshold in either or both race/ethnicity groups indicated by color. Venn diagrams providing pictorial representation of the same comparisons shown to the right of each plot.

A total of 127 traits among EA individuals were associated with genetically-predicted height and measured height at phenome-wide significance (**Figure 2A, Supplemental Table 3**). Thus, we found genetic evidence supporting associations with height for 37% (127/345) of the traits that were associated with measured height. In AA individuals, 2 traits (acquired foot deformities [phecode 735] and dermatophytosis of nail [phecode 110.11]) were associated with genetically-predicted height and measured height at phenome-wide significance (**Figure 2B, Supplemental Table 4**). An additional 46 traits had nominally-significant associations with genetically-predicted height and phenome-wide significant associations with measured height in AA individuals. **Table 2** shows the top ten and closely related phecodes associated with genetically-predicted height in EA individuals that achieve at least nominal significance in AA individuals. Circulatory system was the most frequent phecode group/system among the top ten, consistent with prior studies of height-associated conditions^2-5,14-16,18-20^. In addition, we noted that a number of the conditions in **Table 2** are associated with diabetes mellitus, a condition that is more prevalent among Veterans than in the general population, motivating secondary analyses described below.

**Figure 2.**
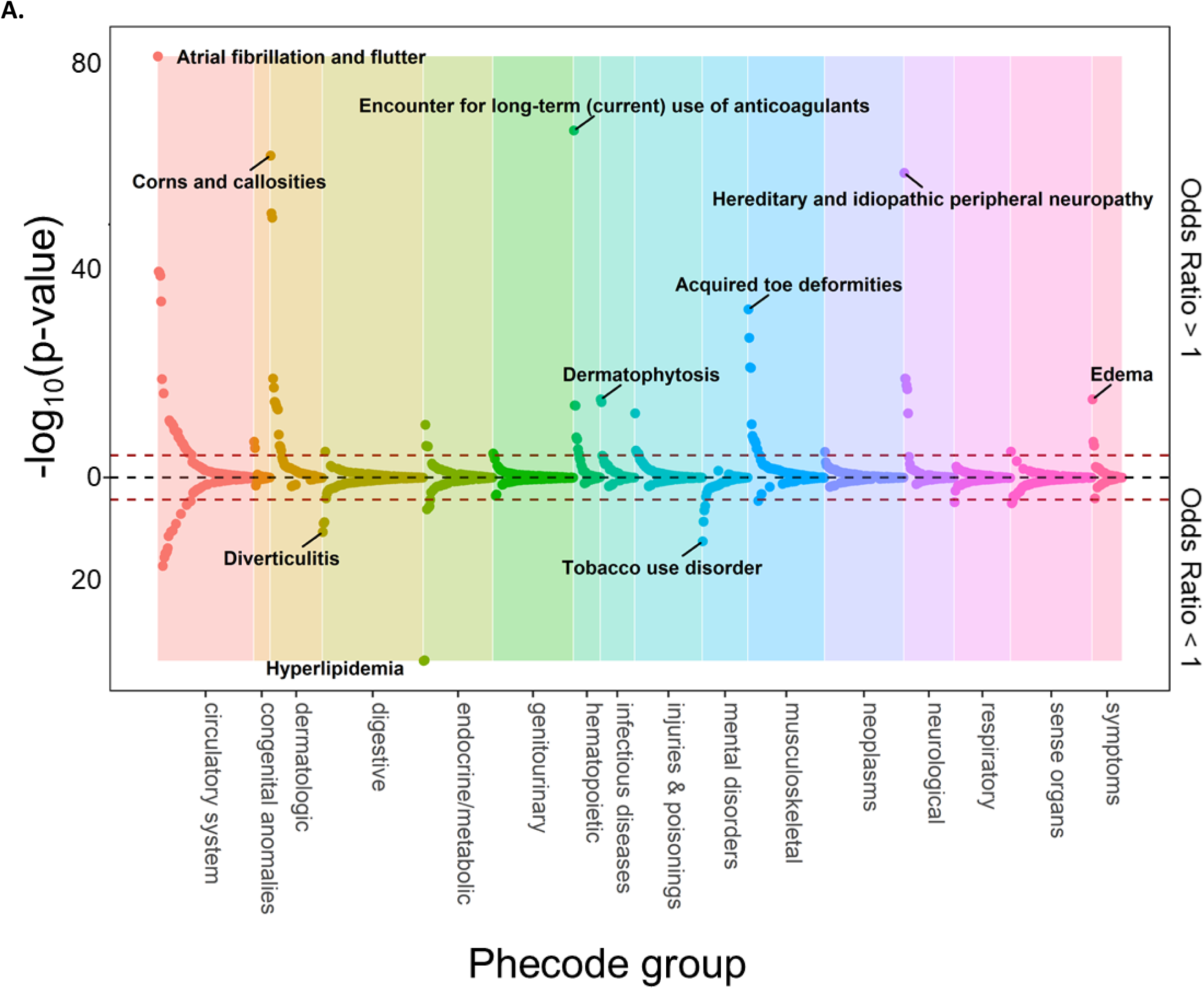
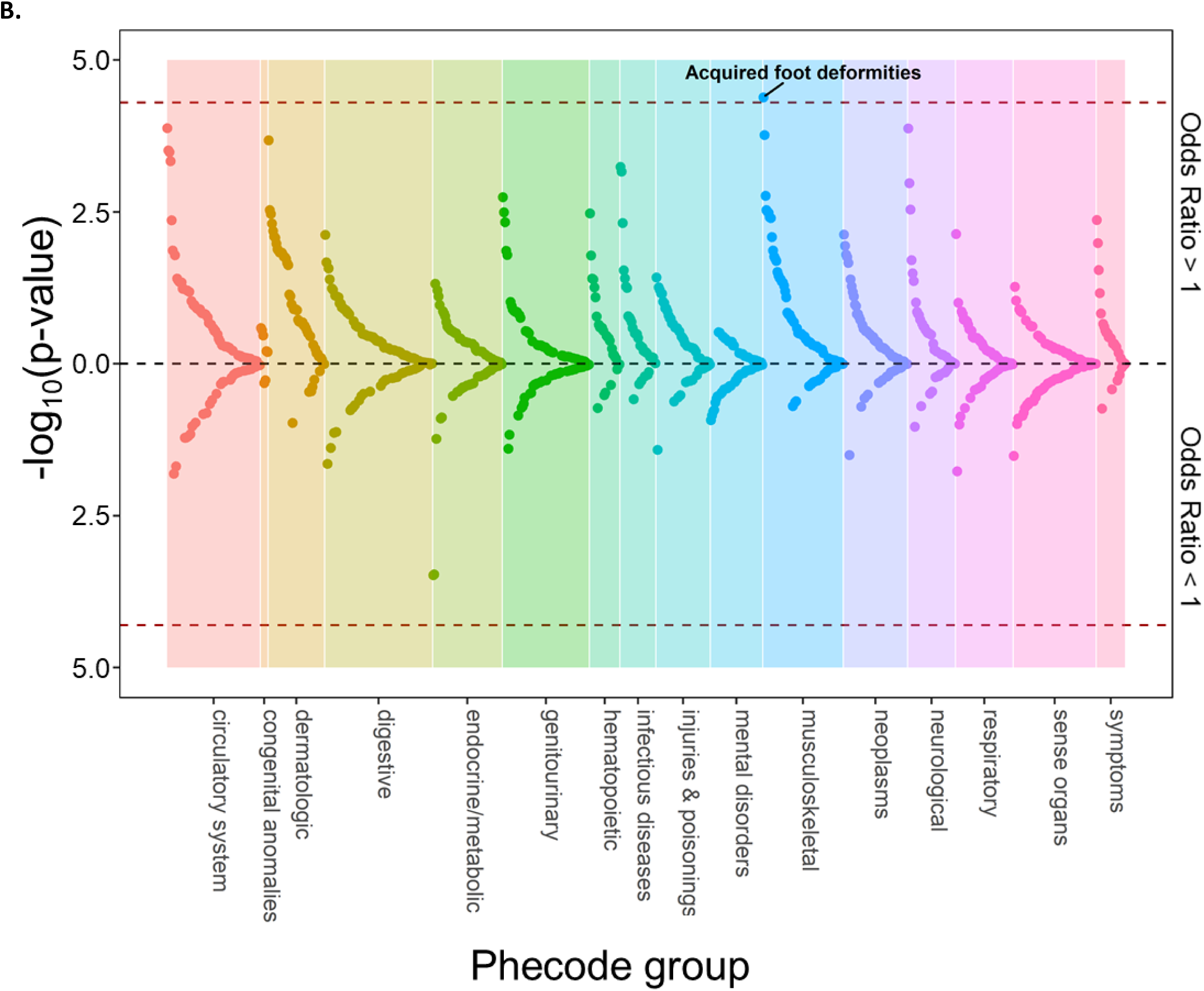
Phenome-wide associations with genetically predicted height in non-Hispanic White (A) and non-Hispanic Black (B) individuals. Plot of phecodes versus -log_10_(p-value) for association with genetically-predicted height in non-Hispanic White (A) and non-Hispanic Black (B) participants in MVP. Phecodes were limited to single decimal place for clarity (e.g., 427.2 for atrial fibrillation or flutter is shown but 427.21 for atrial fibrillation is not). Associations with a negative beta coefficient (i.e., odds ratio < 1) are plotted below the x-axis, and those with a positive beta coefficient (i.e., odds ratio > 1) are plotted above the x-axis. Red dotted lines indicate race/ethnicity-specific phenome-wide significance thresholds (*p* < 3.6E-5 for non-Hispanic White and *p* < 4.8E-5 for non-Hispanic Black). The top association (lowest p-value) within each phecode group is labeled.

**Table 2.**
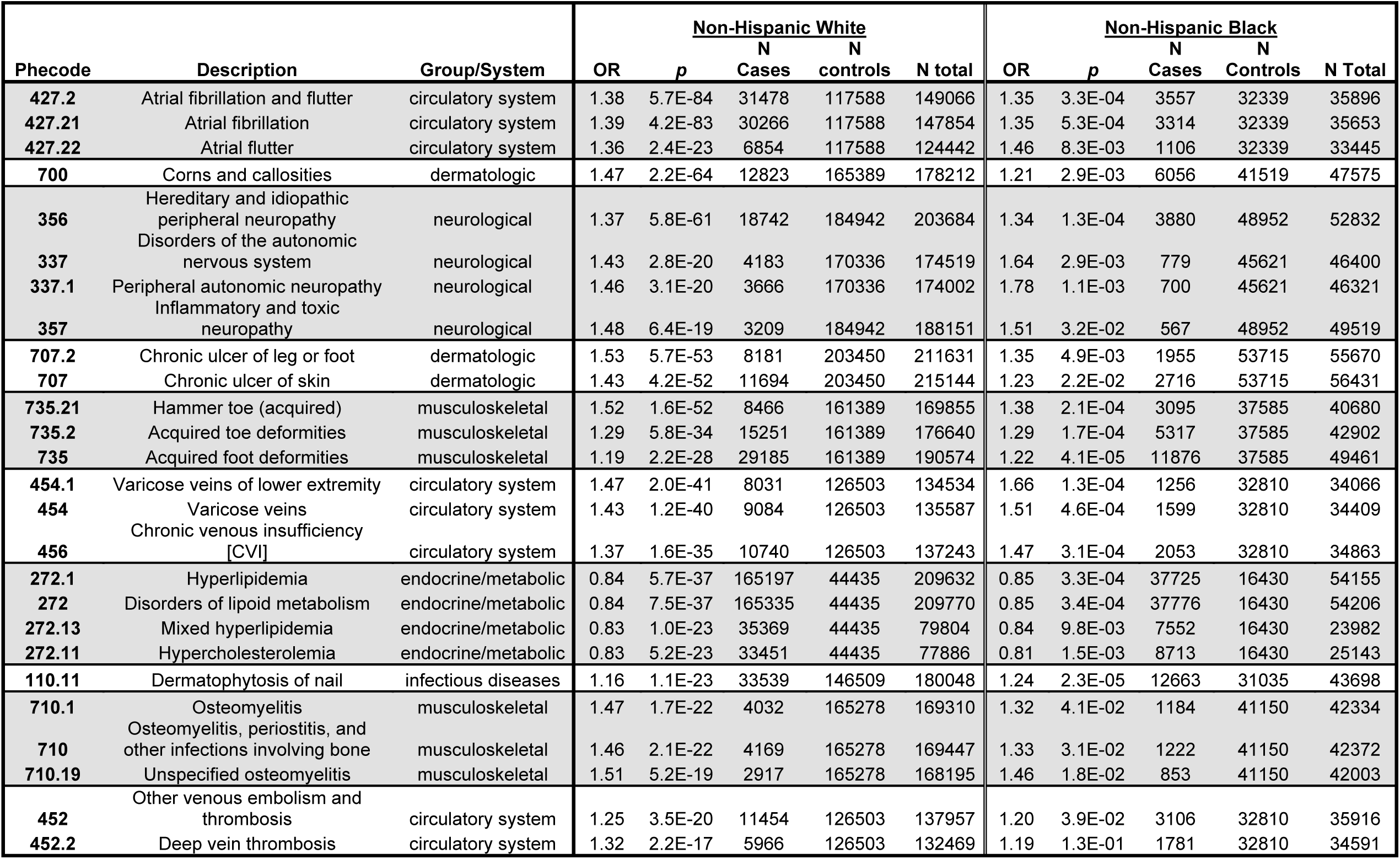
Top phenome-wide associations and closely related traits with genetically predicted height.

BMI is proportional to the inverse of height-squared, and we found that obesity was inversely associated with genetically predicted height (Odds Ratio [OR] 0.94 per SD increase in height, *p*=5.5×10^−7^). To address potential confounding by BMI, we tested whether associations of genetically-predicted height with clinical traits were sensitive to inclusion of BMI as a covariate in the MR-PheWAS analysis. Inclusion of BMI as a coefficient did not substantially impact the beta coefficients for traits associated at phenome-wide significance in EA individuals and at nominal significance in AA individuals (**Supplemental Figure 2**).

### Effect of diabetes mellitus status on genetically-predicted height associations with neurologic, dermatologic, and infectious diabetes complications

In the MR-PheWAS, we found associations of genetically-predicted height with phecodes for several peripheral neuropathy conditions, including diabetic neuropathy (OR 1.18 [95% CI 1.12, 1.24]). However, genetically-predicted height was only modestly associated with diabetes itself with this association not exceeding the phenome-wide significance threshold (**Supplemental Table 5**). To evaluate whether association of genetically-predicted height with peripheral neuropathy was modified by diabetes mellitus, a highly prevalent condition among Veterans^32,33{Affairs, 2015 #159^, we tested the association after stratifying by diabetes mellitus status. The association of genetically-predicted height with hereditary and idiopathic peripheral neuropathy (phecode 356) did not vary by diabetes mellitus status (**Figure 3A**, heterogeneity *p=*0.5). We also noted genetically-predicted height associations with several infectious and dermatologic conditions that are not uncommonly observed in the setting of diabetes and diabetic neuropathy – chronic lower extremity ulcers, osteomyelitis, and superficial cellulitis (**Table 2, Supplemental Table 3**). To determine if genetically-predicted height associations with these dermatologic and infectious complications common in diabetes patients were modified by diabetes mellitus status, we tested the associations stratified by diabetes mellitus. The genetically-predicted height association with chronic leg/foot ulcer (phecode 707.2) was comparable in those without and with diabetes mellitus (heterogeneity *p=*0.6; **Figure 3A**). In contrast, genetically-predicted height associations with osteomyelitis, periostitis and other infections of bone (phecode 710) and with superficial cellulitis and abscess (phecode 681) were stronger in individuals with diabetes mellitus than in those without diabetes mellitus (heterogeneity *p*=3.2×10^−3^ and *p=*2.4×10^−4^ for phecodes 710 and 681, respectively; **Figure 3A**).

**Figure 3.**
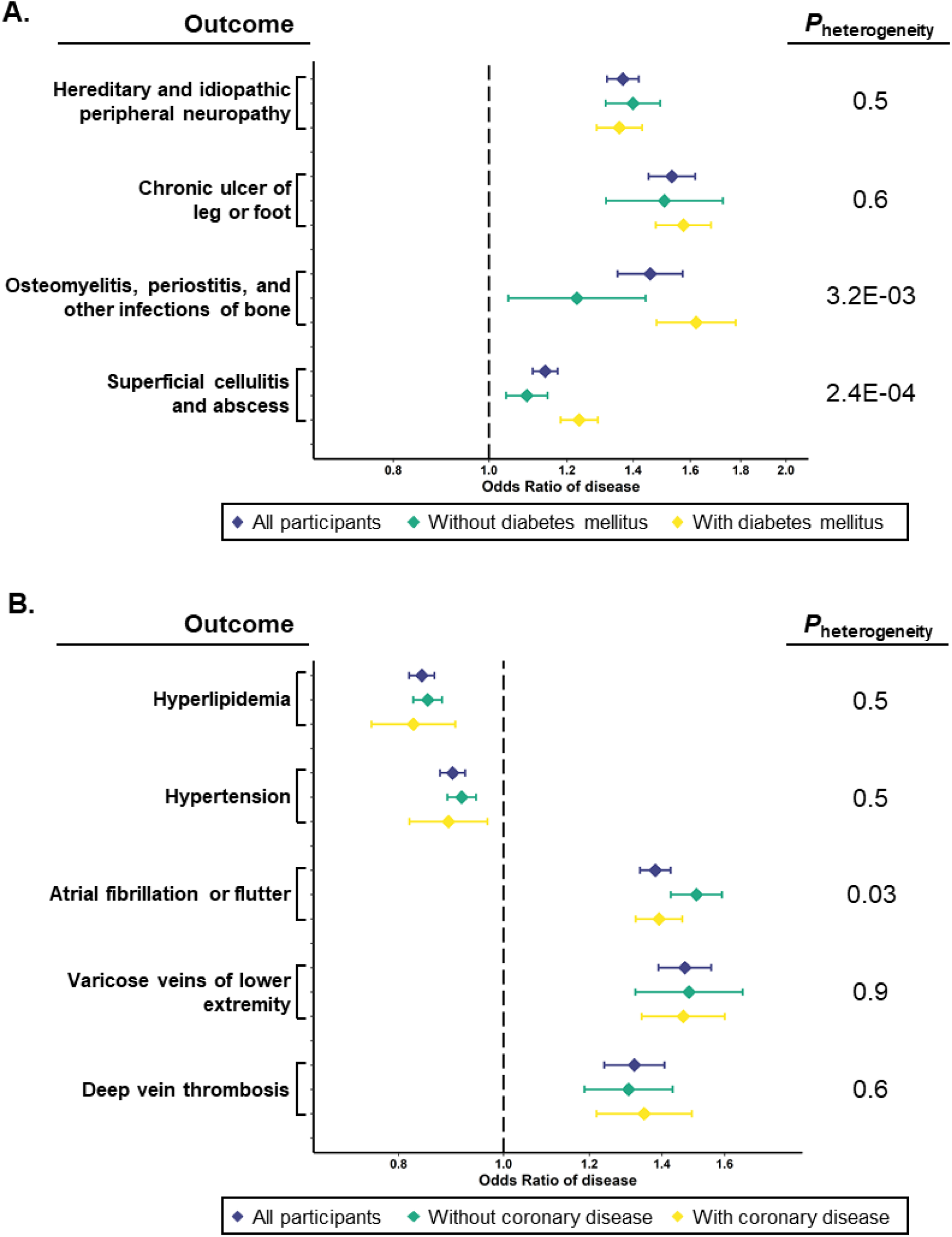
Associations of selected traits with genetically-predicted height after stratifying by diabetes mellitus (A) or coronary heart disease (B) status. Odds ratio (OR) and 95% confidence intervals shown for associations of the indicated traits with genetically-predicted height in all participants (purple), those without diabetes (A, green) or without coronary heart disease (B, green) and those with diabetes (A, yellow) or with coronary heart disease (B, yellow). P-values from test of heterogeneity between strata shown to the right.

### Effect of CHD status on genetically-predicted height associations with CHD risk factors and other circulatory system disorders

Genetically-predicted height has previously been associated with CHD and several CHD risk factors^15,18^, and we reproduced several of these associations (**Supplemental Table 3**). Given that CHD risk factors and CHD are correlated in our clinical data, we examined associations of genetically-predicted height with hyperlipidemia and hypertension stratifying by CHD status. Genetically-predicted height was inversely associated with hyperlipidemia and hypertension in both individuals without and with CHD (heterogeneity *p* >0.05 for both; **Figure 3B**). Atrial fibrillation or flutter is another cardiovascular condition associated with height and genetically-predicted height^5,16,19^. CHD is also a risk factor for atrial fibrillation, but the associations of genetically-predicted height with CHD and atrial fibrillation are in opposing directions. We found that genetically-predicted height was associated with a higher odds ratio for atrial fibrillation or flutter in individuals without CHD (OR 1.51 [95% CI 1.43, 1.59]) than in those with CHD (OR 1.39 [95% CI 1.32, 1.46]; heterogeneity *p*=0.03; **Figure 3B**). Conversely, we did not observe heterogeneity in the associations of genetically-predicted height with varicose veins of the lower extremity or with deep vein thrombosis (heterogeneity *p* >0.05 for both; **Figure 3B**), two other circulatory system disorders associated at phenome-wide significance in the MR-PheWAS.

### Multi-ethnic GWAS of height in the Million Veteran Program

The MR-PheWAS described above was performed using summary statistics from an external GWAS limited to individuals of European ancestry^10^. To determine if a multi-ethnic GWAS in MVP might yield a substantial increase in the number of loci associated with height and thus inform a better instrument for estimating genetically-predicted height particularly in non-European ancestry samples in future studies, we performed GWAS in individuals of non-Hispanic White, non-Hispanic Black, and Hispanic-American race/ethnicity groups in MVP followed by trans-ethnic meta-analysis. **Table 3** summarizes the GWAS results. GWAS in 235,398 EA individuals identified 18 height-associated loci with *p*<5×10^−8^ that had not previously been reported in European-ancestry height GWAS (**Supplemental Table 6**). GWAS in 63,898 AA individuals and 24,497 HA individuals did not identify any novel height-associated loci compared to published European-ancestry GWAS. Trans-ethnic meta-analysis in MVP across all three race/ethnicity groups identified 16 loci associated with height not identified in prior height GWAS in European and African ancestry individuals (**Supplemental Table 7**). Of the 24 variants at the 16 novel loci, only 1 was annotated as a non-synonymous coding variant, and none of the 24 had a scaled CADD score exceeding the 90^th^ percentile, suggesting available genome annotation did not clearly indicate deleterious functional impact of the variants (**Supplemental Table 8**).

**Table 3.** Summary of racen/ethnicity-specific and trans-ethnic meta-analysis genome-wide association study results.

|  | **Known loci** | **MVP EA** | **MVP AA** | **MVP HA** | **EA+AA+HA**<br>**Trans-ethnic**<br>**meta-analysis** |
| --- | --- | --- | --- | --- | --- |
| **Number of loci** | 712 | 533 | 20 | 16 | 488 |
| **Independent SNPs** | 3290 | 2,027 | 20 | 18 | 1,591 |
| **Novel loci** | - | 18 | 0 | 0 | 16 |
| **Independent SNPs**<br>**at novel loci** | - | 22 | 0 | 0 | 24 |

## Discussion

We report associations of genetically-predicted height with clinical traits across a spectrum of systems/domains. To our knowledge the broadest prior MR analysis of height examined 50 traits^15^, and we expand that scope to over 1,000 traits using an MR-PheWAS approach applied to data from the largest integrated healthcare system in the US. We confirm known risk-increasing (atrial fibrillation/flutter) and risk-lowering (CHD, hypertension, hyperlipidemia) associations with cardiovascular conditions and risk factors, as well as recently reported associations with varicose veins^40,41^. In addition, we identified potentially novel associations with peripheral neuropathy and infections of the skin and bones. Although our sample had less statistical power to detect associations in AA individuals compared to EA individuals, we found generally consistent associations of genetically-predicted height with clinical conditions across the two populations. The large sample available through the MVP also permitted analyses stratified by CHD and diabetes mellitus status, revealing heterogeneity in the associations involving atrial fibrillation/flutter and infections common in diabetes patients, respectively. Our results suggest that an individual’s height may warrant consideration as a non-modifiable predictor for a number of common conditions, particularly those affecting peripheral/distal extremities that are most physically impacted by tall stature.

This PheWAS study confirms several associations of cardiovascular and circulatory system disorders with genetically-predicted height that have been previously described in other study samples but also provides some additional insights. First, the effect sizes for height associations with cardiovascular disease traits and risk factors were notably similar between the UK Biobank two-sample study and our one-sample two-stage least squares study in MVP strengthening the weight of evidence supporting causally protective associations between tall stature and hypertension, hyperlipidemia, CHD, and atrial fibrillation. Second, the observation of a stronger association of genetically-predicted height with a lower risk of atrial fibrillation among individuals without CHD is consistent with negative confounding by CHD on the height-atrial fibrillation association. Lastly, we note a complete lack of association between genetically-predicted height and peripheral vascular disease (phecode 443). Whether this finding reflects truly divergent relationships of height with disease in distinct vascular beds warrants future study. The PheWAS results also extend our understanding of the clinical impacts of tall stature beyond cardiovascular disease. Notably, increased stature was disease risk-increasing for the majority of non-cardiovascular conditions, contrary to the pattern of association with cardiovascular disease risk factors and CHD. Two clusters of traits of particular interest were peripheral neuropathy conditions and venous circulatory disorders.

Studies examining risk factors for slowed nerve conduction have previously found an inverse association of height with nerve conduction velocity and amplitude^42,43^. The association of genetically-predicted height with clinical peripheral neuropathy supports the prior epidemiologic findings and suggests that height-related effects on nerve conduction are clinically significant. We also observed associations of genetically-predicted height with extremity complications that are not infrequently observed in the setting of peripheral neuropathy including cellulitis and skin abscesses, chronic leg ulcers, and osteomyelitis. Recent work has found height and peripheral sensory neuropathy to be independent risk factors for diabetic foot ulcer^44,45^. We found consistent associations of genetically-predicted height with peripheral neuropathy and chronic leg ulcer irrespective of diabetes status. In contrast, we observed stronger associations of genetically-predicted height with skin and bone infections in those with diabetes compared to those without diabetes, suggesting synergy between taller stature and other characteristics of diabetes and diabetes care to impact infection risk. To our knowledge, height has not been described as a risk factor for skin and bone infections, in those with or without diabetes, though a plausible mechanism would be via height-related peripheral neuropathy.

Prior observational studies have suggested increased height predisposes to varicose veins and the causality of these associations has been supported through MR analyses^41^. Buttressing these epidemiologic observations are studies demonstrating adverse venous pressure dynamics in taller individuals that likely promote peripheral venous stasis and varicose veins^46^. A second genetic study of varicose veins interestingly found causal associations of circulating levels of varicose vein-related biomarkers with height, suggesting a potential bidirectional causal association between height and varicose veins^40^. In this PheWAS of genetically-predicted height in MVP, we found evidence supporting potentially causal associations of height with varicose veins and venous thromboembolic events and extend that association to a number of other related venous circulatory disorders: chronic venous insufficiency and venous hypertension.

Our multi-ethnicity GWAS in MVP identified 16 height-associated loci that were not found in recent European-ancestry or African-ancestry GWAS meta-analysis. Interestingly, at least two of the loci identified in the trans-ethnic meta-analysis fall near genes – *DLG5* and *SMURF2* – that were not found in conventional GWAS of height in European-ancestry individuals but were identified in analyses that incorporated functional annotation into the association analysis^47^. Another locus, near the *BMP2K*, is highly plausible as a height-associated locus given that its expression is inducible by *BMP2*, a gene that was also associated with height in the aforementioned analysis that incorporated functional annotation into genetic association testing^47^. The *USP44* gene, harboring the only non-synonymous variant identified in this trans-ethnic GWAS meta-analysis, has been associated with type 2 diabetes^48,49^, C-reactive protein levels^50^, and acute myeloid leukemia^51^ in prior GWAS, but has not been implicated in anthropometric traits in prior studies. The trans-ethnic meta-analysis in MVP also replicated one locus on chromosome 10 (near the *ECD* gene) that was identified in a recent African-ancestry GWAS of height^37^ and that had not previously been discovered in European-ancestry GWAS. The identification of additional genetic associations with height in a smaller total sample size than the most recent European-ancestry GWAS supports the importance of non-European populations in characterizing the genetic architecture of complex traits as has been well-described previously^52,53^. Indeed, a multi-cohort, international multi-ancestry height GWAS meta-analysis is nearly complete and will extend the single-study results we report here.

Important limitations to our analyses exist. First, we used loci associated with height from a European-ancestry GWAS meta-analysis to develop the GRS employed in the MR-PheWAS analysis. Thus, the analysis of non-Hispanic Black individuals was limited by a weaker genetic instrument in addition to smaller sample size when compared to the analysis of non-Hispanic White individuals. As multi-ancestry height GWAS meta-analysis are completed, stronger genetic instruments for MR-PheWAS analyses in AA and other non-EA populations may soon be available. Second, we did not interrogate genetic correlation or pleiotropy between height and associated traits identified in the MR-PheWAS. To perform such analyses on a phenome-wide scale exceeds the scope of this manuscript. Thus, we are cautious about causal interpretation of the results of the MR-PheWAS reported here in the absence of such secondary analyses. Given increasing numbers of clinical biobanks globally, replication of MR-PheWAS associations in independent cohorts will be the focus of future work. Third, prior work has demonstrated associations of genetically-estimated height with income and socioeconomic status particularly in men^6^, and we cannot exclude the possibility that associations found in the MR-PheWAS are mediated by socioeconomic status rather than a direct effect of height. Income and education data is available in only a subset MVP of participants, limiting the ability to comprehensively evaluate mediation by socioeconomic variables at the present time. Finally, the sample of individuals receiving care in the US VA Healthcare System may not represent a general US adult population. In particular, US Veterans in this study are mostly older males with higher prevalence of a number of common chronic conditions, including diabetes and cardiovascular disease^32-34^. While the higher burden of disease may make the MVP sample non-representative of a typical adult population, the higher prevalence of many traits enhances statistical power for detecting associations in the PheWAS and MR-PheWAS.

In conclusion, we found genetic evidence supporting associations between height and 127 EHR traits in individuals of non-Hispanic White individuals, 48 of which exhibited nominally-significant associations with genetically-predicted height in non-Hispanic Black individuals.

While much work has focused on inverse associations of genetically-predicted height with CHD and its risk factors, this MR-PheWAS analysis suggests taller stature is associated with higher prevalence of many other clinically relevant traits. In particular, we describe associations of genetically-predicted height with conditions that may result from the effects of increased weight-bearing such as acquired toe and foot deformities, and with peripheral neuropathy traits and venous circulatory disorders, conditions for which epidemiologic and physiologic studies have previously suggested a height-dependence. Finally, we highlight the potential importance of height as a risk factor that can impact the care of common chronic diseases by demonstrating interactions of height with diabetes mellitus on skin and bone infections. Taken together, we conclude that height may be an under recognized non-modifiable risk factor for a wide variety of common clinical conditions that may have implications for risk stratification and disease surveillance.

## Supporting information

STROBE checklist

## Data Availability

All GWAS summary statistics will be deposited in the database of genotypes and phenotypes (dbGaP) at the time of publication in a peer-reviewed journal.

## Acknowledgements

This research is based on data from the Million Veteran Program, Office of Research and Development, Veterans Health Administration, and was supported by awards MVP001 I01-BX004821 and MVP003/028 I01-BX003362. This publication does not represent the views of the Department of Veteran Affairs or the United States Government. We are grateful to our Veterans for their contributions to MVP.

## Author Contributions

Conceptualization: SR, JH, CT, CJO, YVS, TLA

Data Curation: JH, JEH, YAH, HHZ, HZ, SP, ERH

Formal Analysis: SR, JH, CT, EL

Funding Acquisition: JMG, PT, PWFW, KMC, KC, CJO

Methodology: JH, CT, HHZ, HZ, EM, KEN, EL, LAL, BFV, YVS, CJO, TLA

Resources: HZ, JMG, SP, ERH, PT, PWFW, KMC, KC, CJO, YVS, TLA

Supervision: EL, LAL, JMG, PT, PWFW, KMC, KC, CJO

Visualization: SR, CT

Writing – Original Draft Preparation: SR, JH, CT, JEH, CJO, YVS, TLA

Writing – Review & Editing: All authors

