## Supplemental Figures for "Evaluation of height as a disease risk factor through a phenome-wide association study of genetically-predicted height"

**Online Supplemental Figures**

**Supplemental Figure 1.** Effect size comparison of height-associated variants from source European-ancestry GWAS meta-analysis and non-Hispanic White- and non-Hispanic Black MVP participants.

**Supplemental Figure 2.** Effect size comparison of associations of genetically-predicted height with clinical traits in MR-PheWAS without and with body mass index (BMI) as a covariate in non-Hispanic White (A) and non-Hispanic Black (B) individuals.

**Supplemental Figure 1. Effect size comparison of height-associated variants from source European-ancestry GWAS meta-analysis and non-Hispanic White- and non-Hispanic Black MVP participants.**

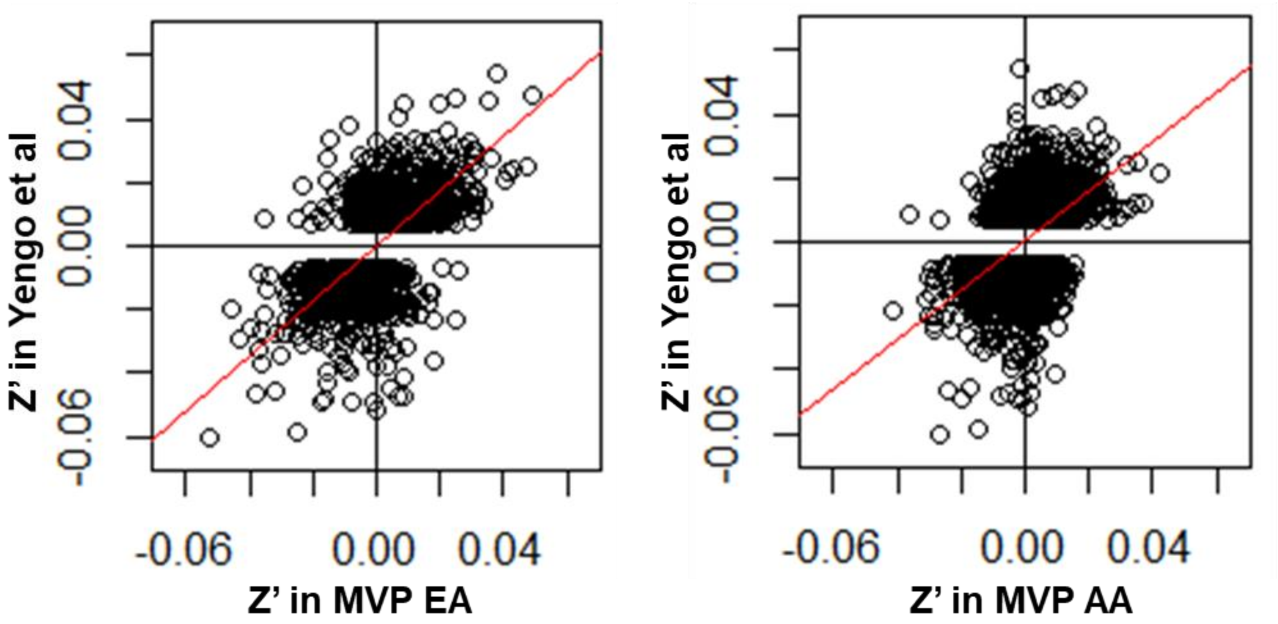


**Slope:** **0.87 [0.84, 0.90]** **0.78 [0.73, 0.82]**

**Supplemental Figure 1**. Comparison of standardized effect sizes (Z’) for associations with height of variants used in height genetic risk score for MR-PheWAS in Yengo et al (*Human Molecular Genetics* 2018:27(20):3641-3649) and MVP non-Hispanic White (EA, left) and non-Hispanic Black (AA, right) individuals.

**Supplemental Figure 2. Effect size comparison of associations of genetically-predicted height with clinical traits in MR-PheWAS without and with body mass index (BMI) as a covariate in non-Hispanic White (A) and non-Hispanic Black (B) individuals.**

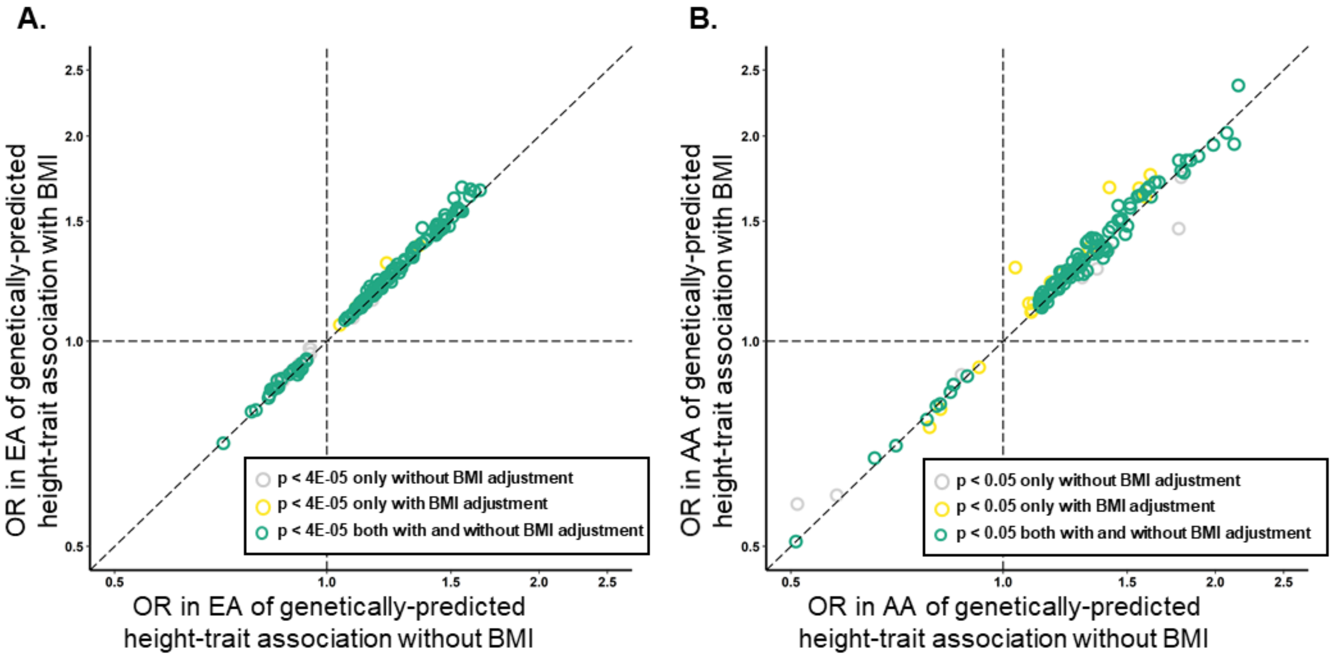
